# Effectiveness of Digital Tools in Promoting Advance Care Planning: A Systematic Review Protocol

**DOI:** 10.1101/2025.07.21.25331958

**Authors:** Ravi Shankar, Fiona Devi, Xu Qian

**Author notes:** **Corresponding Author:** Dr Ravi Shankar; Research and Innovation, Medical Affairs, Alexandra Hospital, Singapore, Email correspondence.

## Abstract

**Background:** Despite compelling evidence supporting advance care planning (ACP) benefits in improving end-of-life care quality and patient autonomy, ACP completion rates remain suboptimal globally, with only 20-40% of adults in developed countries having completed any form of advance directive. Digital health technologies offer promising solutions to overcome traditional barriers to ACP engagement, including accessibility, scalability, and cost-effectiveness challenges. While numerous studies have evaluated individual digital ACP interventions, the evidence base remains fragmented and heterogeneous, with existing reviews often focusing on specific populations, settings, or technologies, limiting generalizability. This systematic review aims to comprehensively evaluate the effectiveness of digital tools and interventions in promoting advance care planning engagement, completion, and quality across diverse populations and healthcare settings, and identify successful intervention components and implementation strategies.

**Methods:** This systematic literature review will adhere to the Preferred Reporting Items for Systematic Reviews and Meta-Analyses (PRISMA) guidelines. A comprehensive search of eight databases (PubMed, Web of Science, Embase, CINAHL, MEDLINE, The Cochrane Library, PsycINFO, and Scopus) will be conducted from inception to July 2025, along with grey literature sources. The review will include randomized controlled trials, quasi-experimental studies, and observational studies evaluating digital health interventions for ACP in adults aged ≥18 years. The PICOS framework will guide study selection, with interventions including web-based platforms, mobile applications, video-based decision aids, telehealth platforms, and emerging technologies. Primary outcomes include ACP engagement rates, advance directive completion, and quality of ACP discussions. Two independent reviewers will conduct study screening, data extraction using Covidence software, and quality appraisal using the Cochrane Risk of Bias tool 2.0 (RoB 2) for randomized trials and ROBINS-I for non-randomized studies. Both narrative synthesis and meta-analysis will be employed where appropriate.

**Discussion:** This protocol outlines a comprehensive systematic review utilizing rigorous methodology to synthesize evidence on digital tools for advance care planning. The findings will provide valuable insights into the effectiveness of various digital ACP interventions, identify optimal intervention components, and highlight implementation considerations. Results will inform healthcare providers, policymakers, and technology developers in implementing evidence-based digital ACP interventions while identifying research gaps requiring future investigation. The review will contribute to ensuring that all individuals have access to effective, culturally appropriate tools that empower them to engage in advance care planning according to their values and preferences.

## Introduction

Advance care planning represents a fundamental component of person-centered healthcare, encompassing the process through which individuals contemplate, discuss, and document their values, goals, and preferences regarding future medical care, particularly in circumstances where they may lose decision-making capacity (1). The conceptual framework of ACP has evolved significantly over the past three decades, transitioning from a narrow focus on completing advance directive documents to a more comprehensive, ongoing process of communication and shared decision-making involving patients, families, and healthcare providers (2). This evolution reflects a growing understanding that effective ACP extends beyond legal documentation to encompass meaningful conversations about values, goals, and preferences that can guide future healthcare decisions.

The importance of ACP is well-established in the literature, with substantial evidence demonstrating its positive impact on multiple outcomes. Research has consistently shown that ACP improves quality of end-of-life care, increases concordance between patient preferences and care received, reduces healthcare utilization and costs, decreases family burden and decisional conflict, and enhances patient and family satisfaction with care (3, 4). Furthermore, ACP has been associated with reduced rates of in-hospital deaths, decreased intensive care unit admissions, and lower likelihood of receiving aggressive life-sustaining treatments at the end of life (5, 6). These benefits extend beyond the individual patient to encompass family members and caregivers, who report lower levels of stress, anxiety, and depression when ACP has occurred.

Despite compelling evidence supporting its benefits, ACP engagement remains disappointingly low across diverse populations and healthcare settings. Studies indicate that only 20-40% of adults in developed countries have completed any form of advance directive, with rates varying significantly by age, ethnicity, socioeconomic status, health literacy, and geographic location (7, 8). Multiple barriers contribute to suboptimal ACP engagement, operating at individual, interpersonal, institutional, and societal levels. Individual-level barriers include lack of awareness about ACP, difficulty understanding complex medical terminology, emotional discomfort discussing death and dying, procrastination, and cultural or religious beliefs that may conflict with planning for end-of-life care (9, 10). Many individuals report not knowing how to initiate ACP conversations or where to access appropriate resources and support.

Interpersonal barriers encompass family disagreement, reluctance to burden loved ones, and communication challenges between patients and healthcare providers (11). Healthcare providers often report feeling inadequately prepared to facilitate ACP discussions, lacking time during clinical encounters, and experiencing their own discomfort with end-of-life conversations. Institutional barriers include time constraints in clinical encounters, lack of systematic approaches to ACP, inadequate provider training, and absence of reimbursement mechanisms (12). Many healthcare systems lack standardized processes for initiating, documenting, and updating ACP preferences, leading to inconsistent practices across providers and settings. Societal barriers involve cultural taboos surrounding death, health disparities, and limited access to healthcare services (13). These multilevel barriers interact in complex ways, creating significant challenges for implementing effective ACP interventions.

The rapid advancement and widespread adoption of digital health technologies present unprecedented opportunities to address many traditional barriers to ACP engagement. Digital tools offer several potential advantages over conventional paper-based or face-to-face ACP approaches. Digital platforms enable individuals to engage with ACP materials at their own pace, in comfortable settings, and at convenient times, removing geographical and temporal constraints associated with clinic-based interventions (14). This flexibility is particularly important for individuals with mobility limitations, those living in rural areas, or those with demanding work or caregiving responsibilities. Once developed, digital interventions can be disseminated to large populations with minimal incremental costs, potentially reaching underserved communities with limited access to specialized ACP facilitation (15).

Digital tools can ensure consistent delivery of evidence-based ACP content, reducing variability in intervention quality and completeness (16). Advanced digital platforms can tailor content based on individual characteristics, preferences, values, and health conditions, potentially improving relevance and engagement (17). The integration of videos, animations, interactive decision aids, and other multimedia elements can enhance understanding of complex medical concepts and facilitate values clarification (18). Digital systems can streamline the documentation, storage, retrieval, and sharing of ACP preferences across healthcare settings and with designated decision-makers (19). While initial development costs may be substantial, digital interventions can potentially reduce long-term costs associated with ACP facilitation and end-of-life care (20).

The landscape of digital tools for ACP has expanded dramatically over the past decade, encompassing diverse technologies and approaches. Web-based platforms offer comprehensive websites providing ACP education, values clarification exercises, advance directive creation tools, and document storage capabilities. Examples include PREPARE (prepareforyourcare.org), MyDirectives, and Vital Talk, which have demonstrated effectiveness in increasing ACP engagement among diverse populations (16, 21). Mobile applications provide smartphone and tablet applications offering portable access to ACP resources, reminders, and communication tools. These include apps like Hello, ALCO, and End-of-Life Planning, which leverage the ubiquity of mobile devices to make ACP more accessible (22).

Video-based decision aids represent another important category of digital ACP tools, providing educational videos that demonstrate medical interventions, explain treatment options, and facilitate informed decision-making about end-of-life care preferences (17). Interactive multimedia programs combine education, values clarification, and decision support through engaging interfaces that guide users through the ACP process (15). Telehealth platforms enable virtual consultation systems allowing remote ACP discussions with healthcare providers or trained facilitators, particularly valuable during the COVID-19 pandemic (23). Electronic health record integration systems incorporate ACP documentation directly into electronic medical records, ensuring accessibility across care settings (24). Emerging technologies include social media platforms leveraging social networking to raise awareness and provide peer support, as well as artificial intelligence and chatbots using natural language processing to guide users through ACP conversations (25, 26).

While numerous studies have evaluated individual digital ACP interventions, the evidence base remains fragmented and heterogeneous. Previous systematic reviews have examined specific aspects of digital ACP tools but have significant limitations. Existing reviews often concentrate on specific populations, settings, or technologies, limiting generalizability (27-30). The rapid pace of technological advancement means that reviews conducted even a few years ago may not capture current innovations and evidence, such as novel app-based or telehealth-supported ACP interventions (31, 32). Many reviews focus primarily on ACP documentation rates without examining broader outcomes such as quality of ACP discussions, goal-concordant care, or patient-reported outcomes (33). Previous reviews often neglect implementation considerations, including user engagement, technology adoption barriers, and sustainability factors (34). Few reviews directly compare different types of digital interventions or identify optimal intervention components (35, 36).

This systematic review aims to address these evidence gaps by comprehensively evaluating the effectiveness of digital tools and interventions in promoting advance care planning engagement, completion, and quality across diverse populations and healthcare settings. The review will identify and categorize different types of digital ACP interventions and their key components, assess the impact of digital ACP tools on patient-reported outcomes, healthcare utilization, and care concordance, examine factors influencing the effectiveness of digital ACP interventions, evaluate the cost-effectiveness of digital ACP interventions compared to usual care or traditional approaches, identify barriers and facilitators to implementing digital ACP tools in various healthcare contexts, and highlight evidence gaps while providing recommendations for future research and practice.

## Methods

This systematic review will be conducted in accordance with the Preferred Reporting Items for Systematic Reviews and Meta-Analyses (PRISMA) guidelines (37) and the Cochrane Handbook for Systematic Reviews of Interventions (38). The protocol has been developed following the PRISMA-P (Preferred Reporting Items for Systematic Review and Meta-Analysis Protocols) statement (39) and has been registered with the International Prospective Register of Systematic Reviews (PROSPERO registration number: CRD420251108596) prior to commencing the review. The systematic approach ensures transparency, reproducibility, and methodological rigor throughout the review process.

### PICOS Framework

The review will employ the PICOS (Population, Intervention, Comparison, Outcomes, Study design) framework to structure the research question and guide study selection. The population includes adults aged 18 years or older, regardless of health status, diagnosis, or care setting, encompassing healthy adults, individuals with chronic conditions, those with serious illnesses, and people at end of life. Studies involving healthcare providers, caregivers, or family members as primary participants will be included if outcomes relate to patient ACP engagement. The intervention comprises any digital health intervention designed to facilitate, promote, or support advance care planning, delivered primarily through electronic or computerized means. The comparison includes studies with any comparator such as usual care, no intervention control groups, alternative ACP interventions, different versions of digital interventions, or pre-post comparisons. Outcomes encompass both primary outcomes (ACP engagement and quality of ACP) and secondary outcomes (patient-reported outcomes, healthcare utilization, end-of-life care outcomes, and implementation outcomes). Study designs include randomized controlled trials, quasi-experimental studies, observational studies with comparison groups, and pre-post studies with adequate sample sizes.

### Search Strategy

The comprehensive search strategy will be developed in collaboration with an experienced medical librarian to ensure optimal sensitivity and specificity. The following electronic databases will be searched from their inception to July 2025: PubMed, Web of Science, Embase, CINAHL, MEDLINE, The Cochrane Library, PsycINFO, and Scopus. These databases were selected to ensure comprehensive coverage of medical, nursing, psychological, and interdisciplinary literature relevant to digital health and advance care planning.

The search strategy combines controlled vocabulary and free-text terms adapted for each database’s specific syntax and indexing system. The search string follows this structure: ((‘digital health’ OR ‘electronic health’ OR ‘eHealth’ OR ‘mHealth’ OR ‘mobile health’ OR ‘web-based’ OR ‘internet-based’ OR ‘online’ OR ‘computer-based’ OR ‘smartphone’ OR ‘mobile application’ OR ‘app’ OR ‘telehealth’ OR ‘telemedicine’ OR ‘video-based’ OR ‘multimedia’ OR ‘virtual’ OR ‘artificial intelligence’ OR ‘chatbot’ OR ‘decision support system*’ OR ‘electronic health record*’) AND (‘advance care planning’ OR ‘advance directive*’ OR ‘living will*’ OR ‘healthcare proxy’ OR ‘durable power of attorney’ OR ‘end-of-life planning’ OR ‘end-of-life care’ OR ‘goals of care’ OR ‘care preference*’ OR ‘treatment preference*’ OR ‘POLST’ OR ‘MOLST’ OR ‘DNR’ OR ‘do not resuscitate’ OR ‘patient preference*’ OR ‘shared decision making’) AND (‘effect*’ OR ‘efficac*’ OR ‘impact*’ OR ‘outcome*’ OR ‘evaluat*’ OR ‘assess*’ OR ‘engagement’ OR ‘completion’ OR ‘uptake’ OR ‘adoption’ OR ‘satisfaction’ OR ‘quality’ OR ‘utilization’ OR ‘implementation’ OR ‘barrier*’ OR ‘facilitat*’ OR ‘knowledge’ OR ‘awareness’ OR ‘self-efficacy’ OR ‘readiness’ OR ‘concordance’)).

Grey literature sources will include ProQuest Dissertations and Theses Global, OpenGrey, ClinicalTrials.gov, WHO International Clinical Trials Registry Platform, and the first 200 results from Google Scholar. Reference lists of included studies and relevant systematic reviews will be hand-searched, and forward citation searching will be conducted using Web of Science. Recent conference proceedings from palliative care, medical informatics, and digital health conferences will be reviewed for relevant abstracts.

### Eligibility Criteria

Table 1 presents the detailed inclusion and exclusion criteria for study selection:

**Table 1.** Inclusion and Exclusion Criteria.

| **Criterion** | **Inclusion** | **Exclusion** |
| --- | --- | --- |
| **Population** | • Adults aged $\geq 18$ years<br>• Any health status or diagnosis<br>• Any care setting<br>• Studies with healthcare providers/caregivers if patient ACP outcomes reported | • Exclusively pediatric populations ($< 18$ years)<br>• Studies with only provider outcomes without patient-level data |
| **Intervention** | • Digital health interventions for ACP<br>• Web-based platforms<br>• Mobile applications<br>• Computer software<br>• Video interventions<br>• Telehealth platforms<br>• EHR-integrated tools<br>• Virtual reality<br>• Social media interventions<br>• Email/text messaging<br>• Multimedia decision aids<br>• AI/chatbot tools | • Primarily paper-based interventions<br>• Digital tools for data collection only<br>• General health websites without ACP focus<br>• EHR systems without ACP functionality |
| **Comparison** | • Any comparator accepted<br>• Usual care<br>• No intervention<br>• Alternative interventions<br>• Different digital versions<br>• Pre-post comparisons | • No exclusions based on comparator |
| **Outcomes** | • Primary: ACP engagement, quality of ACP<br>• Secondary: Patient-reported outcomes, healthcare utilization, end-of-life care outcomes, implementation outcomes | • Studies without relevant ACP outcomes |
| **Study Design** | • RCTs (individual or cluster)<br>• Quasi-experimental studies<br>• Observational with comparison<br>• Pre-post studies ($n \geq 30$)<br>• Mixed methods (quantitative component) | • Case reports/series ($n < 30$)<br>• Purely qualitative studies<br>• Reviews, editorials, commentaries<br>• Conference abstracts only |
|  |  | • Protocols without results |
| **Language** | • English, Spanish, French, German, Chinese | • Other languages |
| **Publication Date** | • No restrictions | • None |
| **Publication Status** | • Peer-reviewed articles<br>• Grey literature meeting quality criteria | • Non-peer reviewed sources without quality assurance |

### Study Selection

Study selection will be managed using Covidence systematic review software, which facilitates independent screening, automatic deduplication, and tracking of reviewer decisions. The selection process will occur in two phases. During title and abstract screening, two reviewers will independently screen all retrieved citations against the eligibility criteria, classifying studies as “include,” “exclude,” or “unclear.” Disagreements will be resolved through discussion, with consultation of a third reviewer if consensus cannot be reached. Studies marked as “unclear” by either reviewer will automatically proceed to full-text screening to ensure potentially relevant studies are not prematurely excluded.

Full-text screening will involve two reviewers independently assessing complete articles for eligibility. Reviewers will document specific reasons for exclusion using a standardized form with predefined categories such as wrong population, wrong intervention, wrong outcomes, wrong study design, or unable to obtain full text. Inter-rater reliability will be calculated using Cohen’s kappa coefficient, with values above 0.80 considered excellent agreement. Prior to commencing screening, all reviewers will participate in a comprehensive training session covering eligibility criteria, use of Covidence software, and resolution of ambiguous cases. A calibration exercise using 50 randomly selected citations will ensure consistency, with reviewers required to achieve at least 80% agreement before proceeding to independent screening.

### Data Extraction

A standardized data extraction form will be developed and piloted on 5-10 studies to ensure comprehensive capture of relevant information. The form will be refined based on pilot testing results and reviewer feedback. Data extraction will capture study characteristics including author information, publication year, country, study design and methodology, sample size calculations, intervention and follow-up duration, funding sources, and conflicts of interest. Population characteristics will include demographic information such as age, sex/gender distribution, race/ethnicity, education level, socioeconomic status, health status and diagnoses, healthcare setting, geographic location, and baseline ACP experience or knowledge.

Intervention characteristics will be extracted in detail, including type of digital technology used, theoretical or conceptual framework guiding intervention development, specific components and features, duration and intensity of intervention, delivery mode, comparison or control conditions, implementation strategies employed, and any co-interventions provided. Outcome data extraction will encompass all outcome measures and assessment tools used, timing of assessments, results for primary and secondary outcomes with effect sizes and confidence intervals, subgroup analyses, adverse events or unintended consequences, and participant satisfaction or acceptability measures.

Two reviewers will independently extract data from each included study, with disagreements resolved through discussion or third-party arbitration. Study authors will be contacted for missing or unclear information, with up to three email attempts over a four-week period. Multiple publications from the same study will be linked and treated as a single unit of analysis to avoid double-counting of participants and outcomes. Extracted data will be entered into a secure, password-protected database with regular backups to ensure data integrity.

### Risk of Bias Assessment

Risk of bias will be assessed using validated tools appropriate to each study design, ensuring comprehensive evaluation of methodological quality. For randomized controlled trials, the Cochrane Risk of Bias tool 2.0 (RoB 2) will be employed, assessing five domains: bias arising from the randomization process, bias due to deviations from intended interventions, bias due to missing outcome data, bias in measurement of the outcome, and bias in selection of the reported result. Each domain will be rated as low risk, some concerns, or high risk of bias, with an overall risk of bias judgment derived from domain-level assessments.

Non-randomized studies will be evaluated using the Risk Of Bias In Non-randomized Studies of Interventions (ROBINS-I) tool, which assesses seven domains: bias due to confounding, bias in selection of participants into the study, bias in classification of interventions, bias due to deviations from intended interventions, bias due to missing data, bias in measurement of outcomes, and bias in selection of the reported result. Each domain will be judged as low, moderate, serious, or critical risk of bias, with an overall assessment reflecting the highest risk level identified across domains.

Pre-post studies without control groups will be assessed using the National Institutes of Health Quality Assessment Tool for Before-After Studies, which evaluates factors such as clear study objectives, eligibility criteria, participant representativeness, sample size justification, outcome measurement validity, and statistical appropriateness. Two reviewers will independently conduct all risk of bias assessments, with disagreements resolved through consensus discussion or consultation with a third reviewer. Risk of bias assessments will be presented in summary tables and figures, and will inform sensitivity analyses excluding high-risk studies.

### Data Synthesis

Data synthesis will employ both narrative and quantitative approaches as appropriate to the available evidence. All included studies will be summarized descriptively, with study characteristics presented in comprehensive evidence tables. A detailed table will map intervention components to reported outcomes, facilitating identification of potentially effective features. Risk of bias assessments will be visually displayed using traffic light plots or similar graphics to provide clear overview of study quality.

Quantitative synthesis through meta-analysis will be considered when at least three studies report the same outcome measure with sufficient statistical information. Given expected heterogeneity in populations, interventions, and settings, random-effects models will be employed using the DerSimonian and Laird method. Effect sizes will be calculated as risk ratios with 95% confidence intervals for dichotomous outcomes and standardized mean differences for continuous outcomes. Statistical heterogeneity will be assessed using the I^2^ statistic, with values of 25%, 50%, and 75% representing low, moderate, and high heterogeneity respectively. The tau^2^ statistic will quantify between-study variance.

Subgroup analyses will be conducted if sufficient studies are available, examining potential effect modifiers including type of digital intervention (web-based versus mobile versus video-based), population characteristics (age groups, health status, cultural background), intervention intensity and duration, level of facilitation or support provided, and healthcare setting. Sensitivity analyses will assess the robustness of findings by excluding studies at high risk of bias, excluding pre-post studies without control groups, using alternative effect size calculations, and examining the impact of different assumptions about missing data.

For outcomes or interventions not amenable to meta-analysis due to heterogeneity or insufficient data, narrative synthesis will follow the framework developed by Popay and colleagues. This will involve developing a preliminary synthesis of findings, exploring relationships within and between studies, assessing the robustness of the synthesis, and developing a narrative that explains patterns in the data. Vote counting based on direction of effect will be used to identify patterns across studies, with harvest plots providing visual representation of evidence distribution. Thematic analysis will identify common intervention components and implementation strategies associated with effectiveness.

Publication bias will be assessed if ten or more studies are included in any meta-analysis, using funnel plots for visual inspection of asymmetry, Egger’s test for statistical assessment, and trim-and-fill analysis to estimate the potential impact of missing studies. The certainty of evidence for primary outcomes will be evaluated using the Grading of Recommendations Assessment, Development and Evaluation (GRADE) approach, considering five domains that may decrease certainty (risk of bias, inconsistency, indirectness, imprecision, and publication bias) and three domains that may increase certainty (large effect, dose-response gradient, and plausible confounding that would reduce demonstrated effect).

## Discussion

This systematic review protocol outlines a comprehensive approach to synthesizing evidence on the effectiveness of digital tools in promoting advance care planning. The protocol’s strengths lie in its broad scope encompassing various digital modalities, comprehensive assessment of outcomes beyond simple documentation rates, attention to implementation factors and equity considerations, and rigorous methodology following established systematic review guidelines. By employing a systematic and transparent approach, this review will provide valuable insights into the current state of evidence while identifying areas requiring further investigation.

The comprehensive search strategy across multiple databases from inception to July 2025 ensures capture of relevant studies across disciplines and geographic regions. The inclusion of grey literature sources and multiple languages reduces the risk of publication and language bias. The use of Covidence software for study selection and data management enhances efficiency and transparency while maintaining methodological rigor. The dual independent review process for all stages of the review minimizes bias and errors in study selection, data extraction, and quality assessment.

The application of the PICOS framework provides clear structure for defining eligibility criteria and ensures comprehensive consideration of relevant factors. By including diverse study designs beyond randomized controlled trials, the review acknowledges the complexity of evaluating digital health interventions in real-world settings. The detailed data extraction form will capture nuanced information about intervention components, implementation strategies, and contextual factors that may influence effectiveness. This granular approach will facilitate identification of successful intervention elements and optimal implementation conditions.

The use of validated risk of bias tools appropriate to each study design ensures consistent and transparent quality assessment. By employing RoB 2 for randomized trials, ROBINS-I for non-randomized studies, and NIH tools for pre-post studies, the review acknowledges the diverse methodological approaches used in digital health research. The planned sensitivity analyses based on risk of bias assessments will help determine the robustness of findings and identify whether methodological quality influences observed effects.

Several challenges are anticipated in conducting this review. The heterogeneity of digital interventions, ranging from simple educational websites to complex artificial intelligence-driven platforms, may complicate synthesis and comparison. The rapid pace of technological innovation means that some interventions evaluated in older studies may no longer be available or relevant. The review will address this by examining temporal trends and emphasizing findings from recent studies while maintaining historical context. The diversity of outcomes measured across studies, including different ACP engagement metrics, quality indicators, and implementation outcomes, may limit opportunities for quantitative synthesis. The review will employ standardized outcome categories and effect size calculations to facilitate comparison where possible.

The quality of available evidence may vary considerably, with many studies likely to have methodological limitations such as lack of randomization, small sample sizes, short follow-up periods, and high attrition rates. The transparent reporting of quality assessments and careful interpretation of findings in light of methodological limitations will be essential. The potential for publication bias, with positive findings more likely to be published, may lead to overestimation of intervention effectiveness. The comprehensive search strategy, including grey literature, and planned publication bias assessments will help address this concern.

Cultural and contextual factors present important considerations for this review. Most digital ACP research has been conducted in Western, English-speaking countries, potentially limiting generalizability to other cultural contexts where attitudes toward death, dying, and advance planning may differ significantly. The review will explicitly examine the geographic and cultural distribution of included studies and discuss implications for global applicability. Attention to health equity considerations will examine whether digital interventions may exacerbate or alleviate disparities in ACP engagement among vulnerable populations.

This protocol has several limitations that should be acknowledged. The restriction to studies published in five languages, while practical given available translation resources, may exclude relevant research from other linguistic contexts. The focus on adult populations excludes potentially important evidence from pediatric advance care planning, which involves unique considerations and stakeholders. The broad definition of digital interventions, while comprehensive, may result in substantial heterogeneity that complicates synthesis and interpretation. The exclusion of purely qualitative studies, while maintaining focus on effectiveness outcomes, may miss important insights about user experiences and implementation barriers.

The anticipated timeline for completing this systematic review is approximately 12 months from protocol registration. Key milestones include database searching and deduplication (months 1-2), title and abstract screening (months 2-4), full-text screening and data extraction (months 4-8), risk of bias assessment and data synthesis (months 8-10), and manuscript preparation and dissemination (months 10-12). Regular team meetings will ensure adherence to timeline and protocol, with any deviations documented and justified in the final report.

Findings from this systematic review will have important implications for multiple stakeholder groups. For healthcare providers and organizations, evidence on effective digital ACP tools can guide selection and implementation decisions, informing integration into routine care workflows. For policymakers, findings regarding cost-effectiveness and implementation requirements can inform coverage decisions and quality improvement initiatives. For technology developers, identification of effective intervention components and user preferences can guide future innovation. For researchers, evidence gaps identified through this review will highlight priority areas for future investigation.

The dissemination strategy will ensure findings reach relevant audiences through multiple channels. The primary systematic review will be submitted to a high-impact journal in palliative care or digital health. Secondary publications may focus on specific populations, intervention types, or implementation considerations. Conference presentations at international meetings will share findings with clinical and research communities. Stakeholder-specific materials, including policy briefs, practice summaries, and patient education resources, will translate findings for diverse audiences. Digital dissemination through websites, webinars, and social media will maximize reach and accessibility.

This systematic review protocol provides a robust framework for synthesizing evidence on digital tools for advance care planning. By employing rigorous methodology, comprehensive scope, and attention to implementation and equity considerations, the review will generate valuable evidence to guide practice, policy, and research. As healthcare systems increasingly embrace digital transformation, understanding how to effectively leverage technology for this fundamental aspect of person-centered care becomes ever more crucial. The findings will contribute to ensuring that all individuals have access to effective, culturally appropriate tools that empower them to engage in advance care planning according to their values and preferences.

## Data Availability

All data produced in the present study are available upon reasonable request to the corresponding author.

